# Potential Neuroprotective Effects of Natural Anti-NMDAR1 Autoantibodies Against Psychiatric Symptoms Associated with Traumatic Brain Injuries

**DOI:** 10.64898/2026.01.08.26343690

**Authors:** Melonie Vaughn, Dean Acheson, Susan Powell, Kate Yurgil, Caroline Nievergelt, Dewleen Baker, Victoria Risbrough, Xianjin Zhou

## Abstract

**Importance:** Traumatic brain injury (TBI) increases the risk of developing psychiatric symptoms such as post-traumatic stress disorder (PTSD), depression and anxiety, however biological risk and resiliency factors that explain the significant heterogeneity in outcomes are limited. Although 5-10% of the population carries natural autoantibodies to the NMDA receptor (anti-NMDAR1) it is unknown if carrying anti-NMDAR1 autoantibodies modifies risk for psychiatric outcomes after TBI.

**Objective:** Since TBI facilitates infiltration of circulating natural anti-NMDAR1 autoantibodies into the brain, we tested the hypothesis that natural anti-NMDAR1 autoantibody levels in plasma may modify risk for development of psychiatric symptoms after TBI.

**Design, Settings and Participants:** Data were analyzed from 1025 Marine Resiliency Study-II participants, a longitudinal study that included plasma collection and assessments for TBI, PTSD (Clinician Administered PTSD Scale-IV), depression (Beck Depression Inventory-2) and anxiety symptoms (Beck Anxiety Inventory) before and after a combat deployment to Afghanistan (2010-2013). Plasma anti-NMDAR1 autoantibody levels were quantified using a luciferase-based immunnoassay. Outcomes were post-deployment symptoms and the predictor was a continuous or dichotomous measure of anti-NMDAR1 autoantibody level. Covariates included pre-deployment symptoms, deployment history and experiences.

**Results:** TBI (606 with TBI, 419 without TBI) was associated with significantly greater depression, PTSD and anxiety symptoms post-deployment. In individuals with no TBI history, anti-NMDAR1 autoantibody levels were not associated with symptoms. In individuals that endorsed a TBI however, higher pre-deployment plasma levels of natural anti-NMDAR1 autoantibodies were significantly associated with lower predicted post-deployment depression and PTSD symptoms, but not anxiety. In the TBI group, high autoantibody group membership lowered predicted post-deployment CAPS-IV and BDI-2 scores by 22 and 25% respectively (Cohen’s d=0.25-0.32). After deployment, prevalence of moderate-severe depression was significantly lower in participants with high anti-NMDAR1 autoantibodies (.8% [2/256 participants]) compared with participants with low anti-NMDAR1 autoantibodies (3.5% [27/763 participants]), as was prevalence of taking psychotropic medications.

**Conclusions and Relevance:** Natural anti-NMDAR1 autoantibodies may be a “resilience” factor for TBI-associated increases in depression and PTSD symptoms, supporting the hypothesis that natural anti-NMDAR1 autoantibodies could have neuroprotective effects. Mechanistic studies are warranted to understand if plasma natural anti-NMDAR1 autoantibodies reach the CNS to suppress glutamate excitotoxicity associated with TBI.

## Introduction

Exposure to a traumatic brain injury (TBI) increases risk for developing posttraumatic stress disorder (PTSD)^1^, depression^2^, and anxiety^2^ up to 2-4 times. There is significant heterogeneity in individual outcomes after TBI however, suggesting potential contribution of both risk and resiliency factors. Following TBI, a rapid and widespread release of glutamate triggers excitotoxicity through activation of extrasynaptic NMDAR (*N*-methyl-D-aspartate receptor) signaling, contributing to secondary neuronal loss and neurodegeneration^3,4^. Interventions that block NMDAR signaling (e.g. memantine), suppress TBI-associated excitotoxic damage, however they can also disrupt adaptive neuroplasticity^5^. Presence of IgG isotype anti-NMDAR1 autoantibodies, which downregulate NMDAR receptors, was suggested to protect against excitotoxicity^6^ but in rare cases is also associated with anti-NMDAR encephalitis. Circulating natural anti-NMDAR1 autoantibodies have been detected in ∼5–10% of the human population, with prevalence levels depending on assay type^7–10^. Given TBI can facilitate infiltration of circulating natural anti-NMDAR1 autoantibodies into the brain, their presence may modulate TBI-associated outcomes depending on titer levels and isotype^11^.

In contrast to cell-based qualitative assays that indicate only “positive” or “negative” antibody presence, we developed a luciferase-based assay to provide accurate quantitative measurements of natural anti-NMDAR1 autoantibodies in blood^12^. Using this approach, we reported that higher levels of natural anti-NMDAR1 autoantibodies in plasma was associated with reduced cognitive decline in patients with Alzheimer’s Disease^13^. This finding is in line with reports that schizophrenia patients seropositive for anti-NMDAR1 autoantibodies exhibit less severe negative symptoms and higher psychosocial functioning than seronegative patients^14,15^. These reports support the hypothesis that anti-NMDAR1 autoantibodies may modify cognitive and psychiatric symptom burdens in patient populations.

To investigate potential effects of natural anti-NMDAR1 autoantibodies on TBI-associated psychiatric outcomes, we leveraged the Marine Resiliency Study II (MRSII) database and plasma bank. This prospective, longitudinal study was the first to show significant associations between TBI and increased risk for psychiatric symptoms ^1^. Using available data and linked plasma samples, we quantified the levels of natural anti-NMDAR1 autoantibodies in the blood of 1,025 Marines and Navy Corpsman before a combat deployment, and examined their associations with post-deployment PTSD, depression, and anxiety symptoms^16,17^.

## Methods

### Study Design and Population

All data and samples were obtained from three all-male infantry battalions participating in the Marine Resiliency Study II (MRSII)^16,17^. In this study, plasma samples were obtained and neuropsychiatric symptoms were assessed 1-4 weeks prior to, and again 4–6 months following, a seven-month combat deployment to Afghanistan (with the final evaluation occurring October 10, 2013). All participants were pre-screened by the Marine Corp to rule out major physical or mental health disorders that precluded their recruitment and participation in combat deployments. Approval for this study was obtained from the institutional review boards of the University of California, San Diego, the Veterans Affairs San Diego Research Service (IRB: H120090 and H180112), and the Naval Health Research Center. Of the 1377 participants originally consented for the Marine Resiliency Study II at pre-deployment, 1025 had available plasma aliquots at pre-deployment and symptom data before and after deployment. All others were excluded due to missing samples or data. Of the 1025, 17% did not go on a combat deployment however they were included in the models as some did incur a TBI during the periods between the first and second assessment (**Table 1**, **Supplemental Table 1**).

**Table 1.**
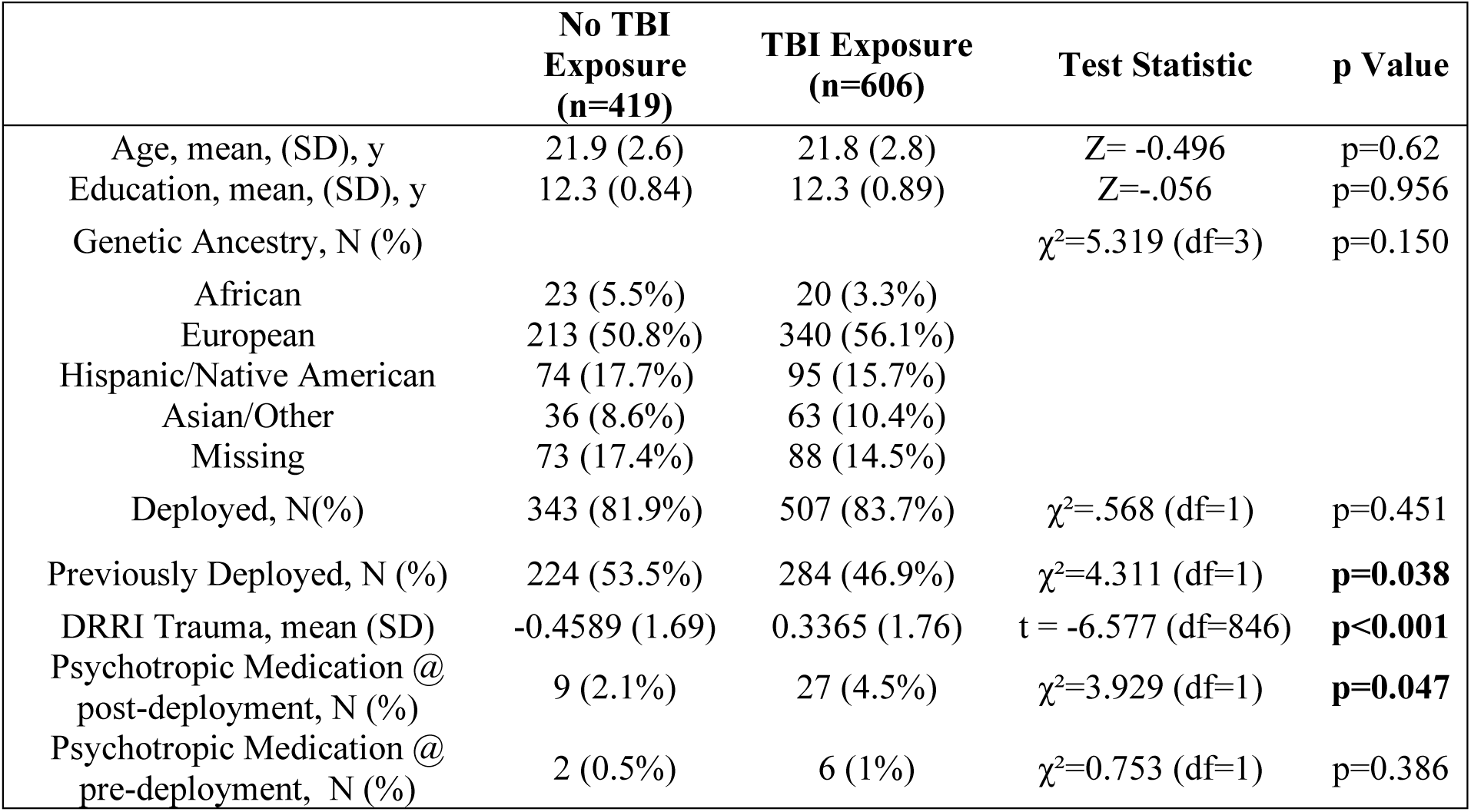
Demographics.

|  | **No TBI**<br>**Exposure**<br>**(n=419)** | **TBI Exposure**<br>**(n=606)** | **Test Statistic** | **p Value** |
| --- | --- | --- | --- | --- |
| Age, mean, (SD), y | 21.9 (2.6) | 21.8 (2.8) | Z= -0.496 | p=0.62 |
| Education, mean, (SD), y | 12.3 (0.84) | 12.3 (0.89) | Z=-.056 | p=0.956 |
| Genetic Ancestry, N (%) | | | $\chi^2=5.319$ (df=3) | p=0.150 |
| African | 23 (5.5%) | 20 (3.3%) |  |  |
| European | 213 (50.8%) | 340 (56.1%) |  |  |
| Hispanic/Native American | 74 (17.7%) | 95 (15.7%) |  |  |
| Asian/Other | 36 (8.6%) | 63 (10.4%) |  |  |
| Missing | 73 (17.4%) | 88 (14.5%) |  |  |
| Deployed, N(%) | 343 (81.9%) | 507 (83.7%) | $\chi^2=.568$ (df=1) | p=0.451 |
| Previously Deployed, N (%) | 224 (53.5%) | 284 (46.9%) | $\chi^2=4.311$ (df=1) | **p=0.038** |
| DRRI Trauma, mean (SD) | -0.4589 (1.69) | 0.3365 (1.76) | t = -6.577 (df=846) | **p<0.001** |
| Psychotropic Medication @<br>post-deployment, N (%) | 9 (2.1%) | 27 (4.5%) | $\chi^2=3.929$ (df=1) | **p=0.047** |
| Psychotropic Medication @<br>pre-deployment, N (%) | 2 (0.5%) | 6 (1%) | $\chi^2=0.753$ (df=1) | p=0.386 |

### Human Plasma Samples

Pre-deployment plasma samples (N=1025) and a subset of post-deployment samples (N=236) from the same individuals were analyzed. Plasma samples had been collected and processed with BD Vacutainer™ Plastic Blood Collection Tubes with Lithium Heparin as described previously^18^. Samples were stored in -80 freezers and kept frozen until immediate use.

### TBI and Symptom Assessments

TBI was assessed via a clinical interview about any lifetime head injuries sustained before the study deployment and injuries sustained between the pre-deployment and post-deployment assessments ^19^. Probable TBI was defined as any head injury resulting in self-reported loss of consciousness (LOC) or altered mental status (i.e., dazed, confused, “seeing stars,” and/or posttraumatic amnesia) immediately afterward or upon regaining consciousness ^19^. Data on timing (endorsement of pre-deployment and/or deployment TBI), type (Blast vs. non-Blast), frequency (i.e. number of injuries) was extracted for exploratory analyses. PTSD symptoms were assessed by a trained clinician using the Clinician-Administered PTSD Scale (0-136, continuous, CAPS-IV) as described^20–22^. Self-reported depression and general anxiety symptoms were collected using the Beck Depression Inventory-II (0-63, continuous, BDI-II)^23^ and Beck Anxiety Inventory (0-63, continuous, BAI) ^24^ respectively. CAPS-IV, BDI-II, and BAI scores were assessed at pre- and post-deployment visits. Endorsement of current psychotropic medication use (positive endorsement for anti-depressants, anti-anxiety, mood stabilizers and/or sleep aids) was also extracted.

### Quantitative Immunoassay

All immunoassays were conducted blinded to demographics and phenotypes. Three positive controls with different levels (high, medium, low) of anti-NMDAR1 autoantibodies (previously established via 10+ rounds of quantification) were included in each batch run and used as references for normalization across 96 well plates (Greiner 96-well Flat Bottom Black Polystyrene plate, Cat. No.: 655097). Goat serum served as negative control and was used for background subtraction. Natural anti-NMDAR1 autoantibodies were quantified in accordance with our previously described method^12^. In brief, 2 µl of human plasma was mixed with protein A/G/L diluted with DAKO antibody diluent (1 µg/µl, Novus Biologicals, NBP2-34985; DAKO, S080983-2), NMDAR1-GLUC fusion protein probe, and 4 µl of diluted goat serum in 1X PBS, 0.25 M NaCl, 1% Triton X100 and incubated for 1.5 hours at room temperature. Samples were washed twice (once in 1X PBS, 0.1% Tween 20, then in 1X PBS) and pelleted by centrifugation (1,889 x g, Eppendorf, Centrifuge 5810R). After resuspending the pellets in 10 µl 1X PBS, 20 µl of Gaussia luciferase substrate (ThermoFisher, cat.16160; Pierce™ Gaussia Luciferase Glow Assay Kit) was added and allowed to stabilize for 10 minutes at room temperature. Luminescence was then quantified using the Tecan Spark plate reader. Relative light units (RLU) for each sample were determined following background subtraction. The ratio of each sample’s RLU to that of the high control was used to rank relative levels of autoantibodies.

### Statistical Analysis

Primary analyses used multiple linear regression models performed via the PROCESS Procedure for SPSS Version 5.0. Documentation available at www.guilford.com/p/hayes. Separate models were estimated for three pre-specified neuropsychiatric outcomes: depression, PTSD, and anxiety. Continuous autoantibody levels were assigned as the key predictor, post-deployment psychiatric scores as the dependent variable, and TBI exposure as the moderator. Pre-deployment symptom scores and deployment status were included as covariates in all models (**Table 2-3**). Interactions with p < 0.10 were probed via simple slopes (**Supplemental Table 2**). All tests were two-sided, and alpha was set to 0.05. Because Bonferroni corrections assume independent tests and these symptoms are highly correlated, this correction would be overly conservative. Therefore, we used false discovery rate (FDR) for multiple-testing correction. Both raw and FDR-adjusted p-values were presented in the results. When a significant interaction between TBI exposure and autoantibody levels was identified in the primary model, follow-up sensitivity analyses were conducted to examine interactions between autoantibody levels and different sub-categories of TBI (deployment, TBI Type, TBI Number). FDR correction was applied within each set of follow-up tests.

**Table 2:** Results from continuous multiple linear regression models. P values <0.05 are bolded. β = Standardized Coefficient (beta), B = unstandardized coefficient, SE = standard error (unstandardized), t = t statistic, CI = confidence interval, AAB: NMDAR1 Autoantibody (continuous autoantibody levels).

| Outcome | Predictor | $\beta$ | B | SE | t value | p value | 95% CI |
| --- | --- | --- | --- | --- | --- | --- | --- |
| PTSD (CAPS) | Intercept |  | 1.2698 | 0.1809 | 7.0186 | <0.0001 | [.9148, 1.6249] |
|  | Pre-Deployment CAPS | 0.0452 | 0.5246 | 0.0293 | 17.9104 | **<0.0001** | [.4671, .5820] |
|  | Deployment Status | 0.4803 | 0.2641 | 0.1577 | 1.6741 | 0.0944 | [-.0455, .5736] |
|  | AAB | 0.0658 | 0.9626 | 0.6631 | 1.4516 | 0.1469 | [-.3386, 2.2638] |
|  | TBI Exposure | 0.0992 | 0.4443 | 0.122 | 3.641 | **0.0003** | [.2048, .6837] |
|  | TBI*AAB | 0.1054 | -1.9146 | 0.8226 | -2.3275 | **0.0201** | [-3.5288, -.3004] |
| | Predictor | $\beta$ | B | SE | t value | p value | 95% CI |
| Depression (BDI-2) | Intercept |  | 0.5417 | 0.1132 | 4.7866 | <0.0001 | [.3196, .7638] |
|  | Pre-Deployment BDI | 0.5302 | 0.5282 | 0.0263 | 20.0689 | **<0.0001** | [.4765, .5798] |
|  | Deployment Status | 0.031 | 0.1155 | 0.0985 | 1.1729 | 0.2411 | [-.0778, .3089] |
|  | AAB | 0.0367 | 0.3421 | 0.4155 | 0.8233 | 0.4105 | [-.4733, 1.1574] |
|  | TBI Exposure | 0.0824 | 0.2356 | 0.0752 | 3.1343 | **0.0018** | [.0881, .3831] |
|  | TBI*AAB | 0.1332 | -1.5388 | 0.5143 | -2.9922 | **0.0028** | [-2.5479, -.5296] |
| | Predictor | $\beta$ | B | SE | t value | p value | 95% CI |
| Anxiety (BAI) | Intercept |  | 0.5344 | 0.1189 | 4.4947 | **<0.0001** | [.3011, .7677] |
|  | Pre-Deployment BAI | 0.4003 | 0.397 | 0.0282 | 14.0717 | **<0.0001** | [.3416, .4523] |
|  | Deployment Status | 0.0413 | -0.1583 | 0.109 | -1.4522 | 0.1468 | [-.3723, .0566] |
|  | AAB | 0.0106 | -0.1534 | 0.4571 | -0.3355 | 0.7373 | [-1.0504, .7437] |
|  | TBI Exposure | 0.1341 | 0.3923 | 0.0831 | 4.7178 | **<0.0001** | [.2291, .5554] |
|  | TBI*AAB | 0.0224 | -0.265 | 0.5668 | -0.4676 | 0.6402 | [-1.3772, .8472] |

Dependent outcome measures were examined for violations of normality assumptions, and predictors were examined for linearity. Post-deployment outcomes of CAPS (PTSD), BDI-II (depression), and BAI (anxiety) measures were heavily rightward skewed as expected for a naturalistic study of symptom development in an initially healthy population. Square root transformations of continuous outcome variables improved model fit, reduced skewness and kurtosis values (< |1|) for both outcomes and their residuals, and enabled regression analyses without violating key statistical assumptions. Autoantibody levels were square root transformed to enhance linearity with the outcomes, improve residual normality, and optimize model fit; this also improved the normality of the predictor distribution. Box-Cox transformation was also evaluated and produced comparable results, supporting the robustness of our findings. To simplify interpretations, only untransformed models are visually presented, along with statistical results from square root transforms.

Since antibody measurement units are not clinically intuitive and may not necessarily exhibit a strong linear relationship with symptom levels (e.g. threshold antibody level may be required for effects), we also converted the continuous autoantibody levels into a categorical factor to validate and help interpret clinical outcome findings. As described in our previous study in a separate population ^13^, participants falling in the top quartile for autoantibody levels (n = 257) were labeled as the “High” group, while those in the lower 75% (n = 768) were designated as the “Low” group. ANCOVA incorporating Type III ANOVA was conducted to examine pre-specified interactions between categorical autoantibody group and TBI exposure. All tests were two sided, and α was set to 0.05. As the comparisons of interest were defined *a priori*, (antibody group comparisons limited to TBI group); direct pairwise comparisons were conducted with Bonferroni correction applied where appropriate. Effect sizes were reported as Cohen’s d. Thus, significant findings from continuous autoantibody models were corroborated by categorical analyses and visualizations.

Several potential covariates were considered for inclusion in the primary multiple linear regression analyses: deployment trauma, genetic ancestry, and previous deployment history. To account for variability in traumatic experiences during deployment, we utilized post-deployment scores from the modified 16-item, 5-point Likert version of the Deployment Risk and Resilience Inventory-2 (DRRI-2) ^25^ Combat Experience and Post-Battle Experience subscales. Scale scores were converted to Z scores and then added together to create a composite combat trauma variable. MRS-II participants were previously genotyped for genetic ancestry, with estimates resulting in four key groups: African, European, Hispanic/Native American, and Asian/Other ^26^. Subject ancestry was dummy coded (0/1) for each of the 4 ancestral groups and put into our primary models to identify influences on our interaction of interest. With nearly half of all participants reported undergoing a previous deployment before enrollment in the present study (508, 49.6%), we examined pre-deployment status as a potential covariate.

Including genetic ancestry, previous deployment history, and trauma exposure as covariates did not meaningfully change the estimated effect of the TBI by autoantibody interaction compared to our base model (all Δβ <0.019; all significant interaction p-values remained <0.03 after corrections). This result was consistent when adding all covariates individually or in unison. Since the primary goal of our analyses is to estimate moderation effect and avoid overfitting, we kept covariates that were theoretically justified and materially influenced the relationship between primary predictor and outcome (pre-deployment symptom scores and deployment status).

For demographic data, comparisons between TBI and No TBI groups were conducted via Mann Whitney U or two-sided t-test as appropriate. Categorical data was examined by performing chi square. Pearson’s correlation was conducted to assess stability of plasma natural anti-NMDAR1 autoantibodies between pre- and post-deployment.

## Results

### Sample Characteristics

Demographics across TBI (N=606, 59.1%) and non-TBI (N=419, 40.9%) groups can be found in **Table 1**. There are no significant differences in age, education, race, deployment status, and previous psychotropic medication use between the groups. DRRI trauma score was significantly higher in the TBI group than the non-TBI group. Within the TBI group, 423 (41.3%) reported only pre-deployment TBIs, 183 (17.9%) reported sustaining a TBI during the deployment period, 152 (14.5%) reported receiving a blast TBI and 350 (34.1%) reported sustaining >1 TBI in their lifetime. The vast majority of TBIs reported were mild in severity (loss of consciousness <30 min, 86%) with only 87 (14%) reporting a moderate to severe TBI (as defined by loss of consciousness >=30 min ^19^).

### Primary Analyses

Pearson’s correlation analysis revealed a strong and statistically significant linear relationship between pre- and post-deployment antibody levels (R=0.83, p < 0.001; **Supplemental Figure 1**), supporting long-term stability of the autoantibodies over the pre- and post-deployment assessment period. There are no significant effect of age, education, ancestry, and previous psychotropic medication on the levels of natural anti-NMDAR1 autoantibodies. Similarly, we found no significant differences in these variables age, education, ancestry, or TBI status when the autoantibody levels were categorized into high and low groups (top 25% vs bottom 75%, see **Supplemental Table 1**). A significantly greater proportion of subjects in the low autoantibody group reported taking psychotropic medications upon returning from deployment (χ² =7.594, df =1, p =0.006), whereas no significant difference was observed prior to deployment (χ² =0.54, df=1, p =0.462).

### Linear Regression Model

Using multiple linear regression models, we tested our primary hypotheses that anti-NMDAR1 autoantibody levels modify TBI exposure effects on post-deployment depression, PTSD, and anxiety symptoms (for linear regression model statistics see **Table 2**). Anti-NMDAR1 autoantibody levels significantly modified TBI associations with depression (**Figure 2A**, **Table 2**) and PTSD (**Figure 2C**, **Table 2**), but not anxiety symptoms (**Table 2**, **Supplemental Figure 2A**). Follow up simple slope analysis (**Supplemental Table 2**) in the TBI group showed significant reductions in predicted post-deployment depression and PTSD symptoms with increased autoantibody level (**Figure 2A, 2C**).

**Figure 1.**
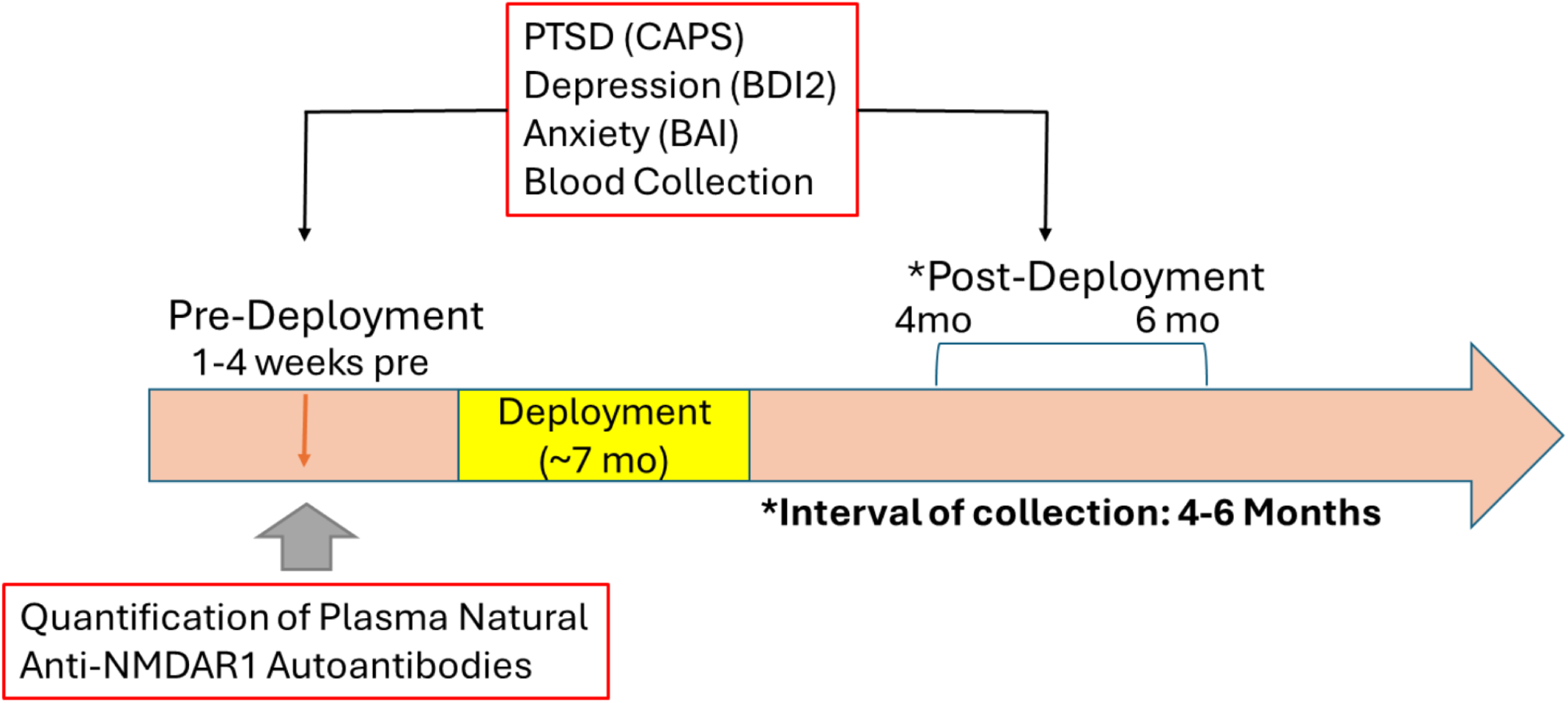
Diagram illustrating blood collection, neuropsychiatric assessment, deployment timeline, and anti-NMDAR1 autoantibody quantification in the MRSII cohort.

**Figure 2.**
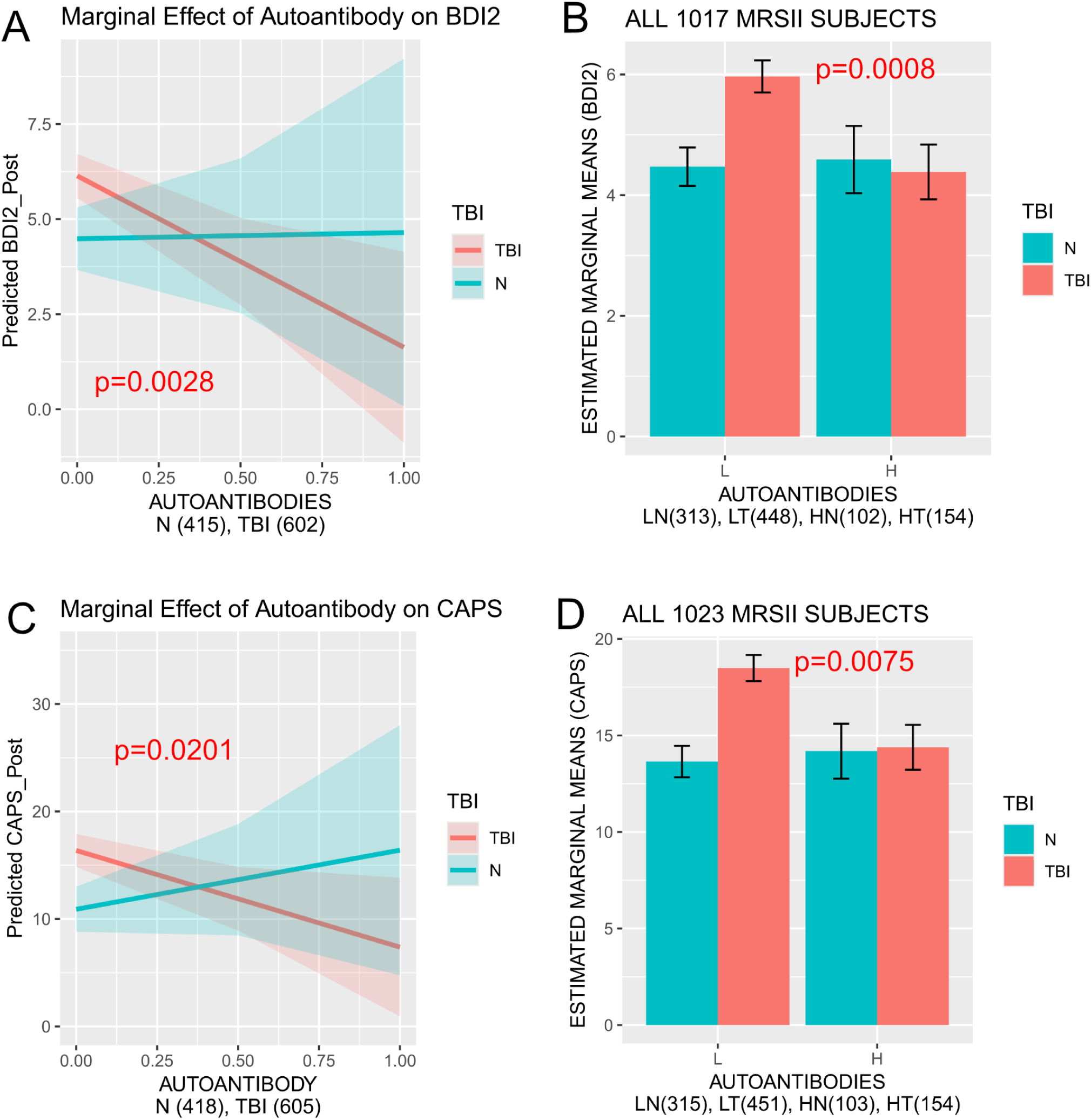
Association of high levels of anti-NMDAR1 autoantibody levels with reduced scores of psychiatric symptoms in the presence of TBI. Legend: N = No TBI Exposure across lifetime, TBI= TBI Exposure across lifetime. All models are visualized using non-transformed data, while statistical values are derived from linear models. P values < 0.10 are reported and color matched with TBI groups on the graphs. (**A)** Effect of continuous autoantibody levels on adjusted BDI2 depression scores. Shaded regions represent the 95% confidence interval. The regression model revealed a significant interaction between autoantibody levels and TBI (Table 2, B = -1.5388, p = 0.0028), consistent with protective effect of autoantibodies in the presence of TBI. (**B**) Interaction between TBI and autoantibody levels as a categorical factor (High autoantibody group is individuals with levels in the top 25%, Low antibody group is bottom 75%) on adjusted BDI2 depression scores. Significant interaction was confirmed (F(1,1010) = 8.5958, p = 0.0034; FDR-adjusted p = 0.0102). Pre-specified comparison between low and high autoantibody groups with TBI (p=0.0008), Error bars = +\- SE. **(C)** Effect of continuous autoantibody levels on adjusted CAPS PTSD scores. Shaded regions represent the 95% confidence interval. The regression model revealed a significant interaction between autoantibody levels and TBI (B = -1.9146, p=0.0201). **(D)** Interaction between TBI and autoantibody levels as a categorical factor on adjusted CAPS PTSD scores. Significant interaction was confirmed (F(1,1017)=5.5491, p=0.0187; FDR-adjusted p=0.0281). Pre-specified comparison between low and high autoantibody groups with TBI (p = 0.0075). Error bars = +\- SE.

### ANCOVA Model

To further validate and interpret the observed interactions, anti-NMDAR1 autoantibody levels were dichotomized into the top 25% of autoantibody levels (“High”) vs. the remaining 75% of the remaining population (“Low”). ANCOVA confirmed significant autoantibody and TBI interactions on depression (F(1,1010)=8.5958, p=0.0034; FDR-adjusted p=0.0102) and PTSD symptoms (F(1,1017)=5.5491, p=0.0187; FDR-adjusted p=0.0281), but not anxiety (F(1,1006)=0.7764, p=0.3784; FDR-adjusted p=0.3784; **Supplemental Figure 2B**). Membership in the high autoantibody group resulted in 22% and 25% lower predicted scores for both depression (p = 0.0008; **Figure 2B**) and PTSD symptoms (p = 0.0075; **Figure 2D**) respectively. Prevalence of moderate-severe depression (BDI-2>19) was significantly lower in participants with high anti-NMDAR1 autoantibodies (.8% [2 of 256 participants]) compared with participants with low anti-NMDAR1 autoantibodies (3.5% [27 of 763 participants]) after deployment (χ² =5.27, p =0.0217). Autoantibody groups did not significantly differ in prevalence for partial PTSD (CAPS-IV>40) or PTSD diagnosis based on the DSM-IV.

### Secondary Analyses

We next conducted a series of follow-up analyses to explore if symptoms associated with different TBI timing, number of TBI injuries or TBI type is differentially modified by autoantibody presence. To examine associations with timing of the TBI in relation to symptom assessment, we partitioned the groups by if they endorsed pre-deployment TBI(s) only, if they endorsed a deployment TBI, or had no TBI history. Both pre-deployment only and deployment TBI groups showed a significant reduction in predicted depression and PTSD symptoms at post-deployment with increasing autoantibody levels (**Table 3**, **Figure 3A-3B**), although effect size of autoantibody modification of PTSD symptoms was the weakest in the deployment TBI group (for simple slopes see **Supplemental Table 2**). Predicted depression and PTSD symptoms were reduced across groups endorsing 1 or >1 TBI injuries (**Figure 3C-3D**, **Supplemental Table 2**). Interestingly, when comparing blast vs. non-blast injuries, anti-NMDAR1 autoantibodies were significantly associated with reduced depression and PTSD symptoms in the non-blast but not blast group (**Figure 3E-3F**, **Supplemental Table 2**). Categorical analyses of high vs. low antibody group generally fit the pattern of results, with depression symptoms being consistent lower independent of TBI timing, injuries or type (**Supplemental Figure 3**). Effect sizes of autoantibodies, treated as a categorical factor, were calculated and summarized for both depression and PTSD symptoms (**Supplemental Table 3**). Predicted PTSD symptoms in blast injury and deployment TBI groups however were not significantly modified by autoantibody group (**Supplemental Figure 3**), although this lack of significance should be interpreted cautiously due to relatively low sample sizes in these groups (N=35-42). Because most blast TBIs occurred during deployment, we investigated whether the reduced effect of autoantibodies on predicted CAPS-IV is attributable to blast exposure or timing of TBI (pre-deployment vs. deployment). Autoantibodies appear to provide comparable effects after blast TBI when it occurs pre-deployment, suggesting that the reduced efficacy of autoantibodies may be more related to when the TBI occurred (i.e. during recent deployment) rather than the TBI type. However, after stratifying TBI into five groups, the sample sizes were insufficient to support definitive conclusions (**Supplemental Figure 4**).

**Figure 3.**
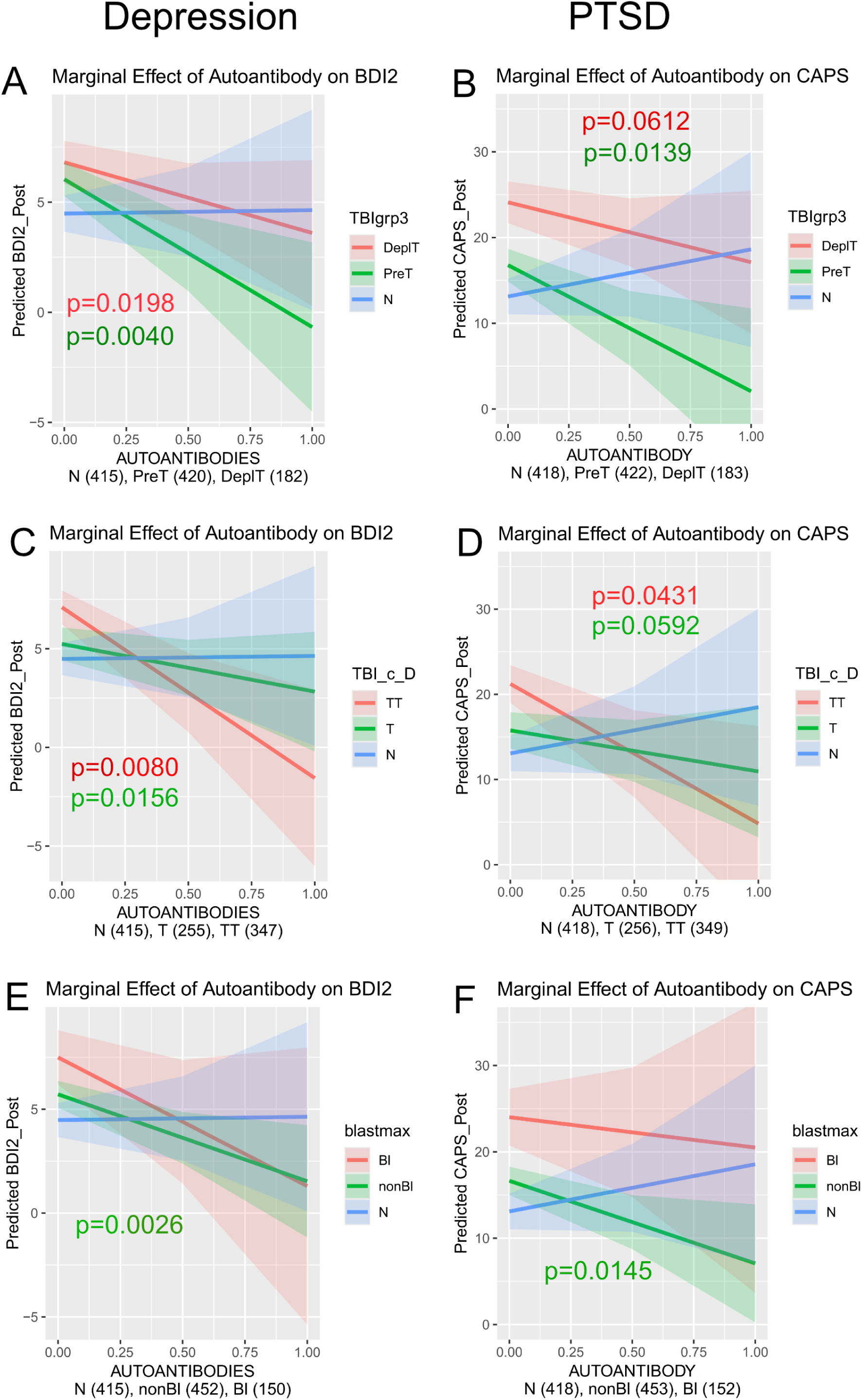
Secondary analyses examined the association between high anti-NMDAR1 autoantibody levels and post-deployment depression and PTSD across TBI timing (A–B), number of injuries (C–D), and injury type (blast vs. non-blast; E–F). All models are visualized using non-transformed data, while statistical values are derived from linear models. P values < 0.10 for simple slopes following significant interactions (see **Table 3**) are reported and color matched with TBI group on the graphs. Shaded regions represent the 95% confidence interval. Sample numbers are presented in order of depression and then PTSD groups. **Panel A-B** TBI timing: TBIgrp3 (TBI groups): N= No TBI Exposure, PreT= Pre-deployment TBI(s) only, DeplT= Deployment TBI. **(A)** Significant autoantibody and TBIgrp3 interactions on adjusted BDI2 depression scores were observed for both pre-deployment (B = –1.6116, p = 0.004) and deployment TBI (B = –1.5425, p = 0.0198). **(B)** Significant autoantibody and TBIgrp3 interactions on adjusted CAPS PTSD scores were observed for pre-deployment (B = – 2.1709, p = 0.0139) and a trend toward significance for deployment TBI (B = –1.9551, p = 0.0612). **Panel C-D** Number of TBI injuries: TBI_c_D (TBI cumulative Deployment): N= No TBI Exposure, T= Single TBI, TT= Multiple TBIs. **(C)** Significant autoantibody and TBI_c_D interactions on adjusted BDI2 depression scores were observed for both single (B = –1.4421, p = 0.0156) and multiple TBI (B = –1.5855, p = 0.008). **(D)** Significant autoantibody and TBI_c_D interaction on adjusted CAPS PTSD scores were observed for multiple TBI (B = –1.9336, p = 0.0431) and a trend toward significance for single TBI (B = –1.799, p = 0.0592). **Panel E-F** TBI Type: blastmax: N= No TBI Exposure, nonBl= Non-blast TBI only, Bl= Blast TBI. **(E)** Significant autoantibody and blastmax interaction on adjusted BDI2 depression scores was observed for non-blast (B = –1.619, p = 0.0026) and a trend toward significance for blast TBI (B = –1.2586, p = 0.1034). **(F)** Significant autoantibody and blastmax interaction on adjusted CAPS PTSD scores was observed for non-blast (β = –2.0802, p = 0.0145).

**Table 3:**
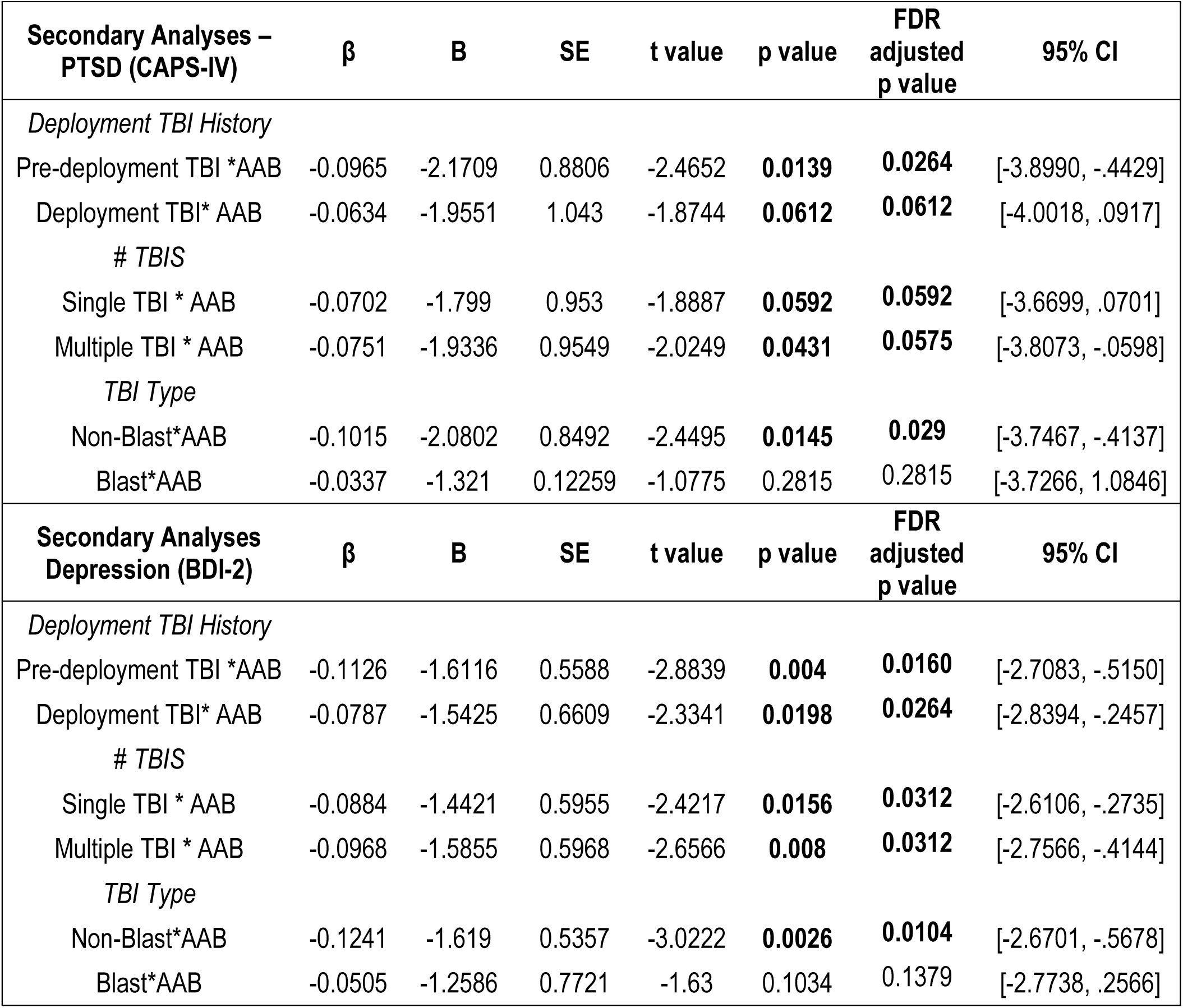
Results from continuous multiple linear regression models. P values <0.10 are bolded. β = Standardized Coefficient (beta), B = unstandardized coefficient, SE = standard error (unstandardized), t = t statistic, CI = confidence interval, AAB = continuous autoantibody levels.

| **Secondary Analyses – PTSD (CAPS-IV)** | **β** | **B** | **SE** | **t value** | **p value** | **FDR adjusted p value** | **95% CI** |
| --- | --- | --- | --- | --- | --- | --- | --- |
| *Deployment TBI History* |  |  |  |  |  |  |  |
| Pre-deployment TBI * AAB | -0.0965 | -2.1709 | 0.8806 | -2.4652 | **0.0139** | **0.0264** | [-3.8990, -.4429] |
| Deployment TBI* AAB | -0.0634 | -1.9551 | 1.043 | -1.8744 | **0.0612** | **0.0612** | [-4.0018, .0917] |
| *# TBIS* |  |  |  |  |  |  |  |
| Single TBI * AAB | -0.0702 | -1.799 | 0.953 | -1.8887 | **0.0592** | **0.0592** | [-3.6699, .0701] |
| Multiple TBI * AAB | -0.0751 | -1.9336 | 0.9549 | -2.0249 | **0.0431** | **0.0575** | [-3.8073, -.0598] |
| *TBI Type* |  |  |  |  |  |  |  |
| Non-Blast* AAB | -0.1015 | -2.0802 | 0.8492 | -2.4495 | **0.0145** | **0.029** | [-3.7467, -.4137] |
| Blast* AAB | -0.0337 | -1.321 | 0.12259 | -1.0775 | 0.2815 | 0.2815 | [-3.7266, 1.0846] |
| **Secondary Analyses Depression (BDI-2)** | **β** | **B** | **SE** | **t value** | **p value** | **FDR adjusted p value** | **95% CI** |
| *Deployment TBI History* |  |  |  |  |  |  |  |
| Pre-deployment TBI * AAB | -0.1126 | -1.6116 | 0.5588 | -2.8839 | **0.004** | **0.0160** | [-2.7083, -.5150] |
| Deployment TBI* AAB | -0.0787 | -1.5425 | 0.6609 | -2.3341 | **0.0198** | **0.0264** | [-2.8394, -.2457] |
| *# TBIS* |  |  |  |  |  |  |  |
| Single TBI * AAB | -0.0884 | -1.4421 | 0.5955 | -2.4217 | **0.0156** | **0.0312** | [-2.6106, -.2735] |
| Multiple TBI * AAB | -0.0968 | -1.5855 | 0.5968 | -2.6566 | **0.008** | **0.0312** | [-2.7566, -.4144] |
| *TBI Type* |  |  |  |  |  |  |  |
| Non-Blast* AAB | -0.1241 | -1.619 | 0.5357 | -3.0222 | **0.0026** | **0.0104** | [-2.6701, -.5678] |
| Blast* AAB | -0.0505 | -1.2586 | 0.7721 | -1.63 | 0.1034 | 0.1379 | [-2.7738, .2566] |

## Discussion

We developed a novel immunoassay to quantify the levels of natural anti-NMDAR1 autoantibodies in blood. Unlike commonly used cell-based assays, our quantitative approach demonstrated that humans carry these autoantibodies to varying degrees. Using this assay, we found that Marines with higher levels of plasma natural anti-NMDAR1 autoantibodies showed significantly lower scores for TBI-associated depression and PTSD symptoms than those with lower levels, consistent with findings from studies on Alzheimer’s disease^13^ and schizophrenia ^14,15^. Given that blood anti-NMDAR1 autoantibodies suppress glutamate excitotoxicity in animal models of stroke and epilepsy ^6^, we suggest that natural anti-NMDAR1 autoantibodies are neuroprotective against TBI-associated neuropsychiatric symptoms. Our analyses using autoantibody levels as a categorical factor suggest that at least 1 quarter of the general population carries high enough levels of these autoantibodies to have functional and clinical significance.

Participants with higher levels of natural anti-NMDAR1 autoantibodies endorsed use of significantly fewer psychotropic medications upon return compared to those with lower levels. In addition, natural anti-NMDAR1 autoantibodies are more strongly associated with reduced depression than PTSD symptoms following TBI, with no significant effect on anxiety. Interestingly, when TBI exposure was stratified into different categories, we found that autoantibodies showed weaker protection against PTSD symptoms in participants with either deployment TBI or blast TBI. Two possible factors may account for this finding. First, the deployment TBI and blast TBI groups were relatively small (42 and 35 subjects, respectively), suggesting that the absence of a significant effect may reflect limited statistical power. Second, delayed-onset PTSD—typically emerging more than six months after trauma—is more common among service members returning from deployment ^27–29^. Autoantibodies may provide stronger protection against chronic or delayed PTSD symptoms than against acute PTSD symptoms, which may account for the observed lower protective effects in deployment TBI.

Ketamine, a non-competitive NMDAR antagonist, is an FDA-approved treatment for depression^30^. Blocking extrasynaptic NMDARs has been hypothesized to contribute to ketamine’s anti-depressant effects^31^. In addition to depression, ketamine also shows promise in treating PTSD^32–34^. These findings align with our observation that natural anti-NMDAR1 autoantibodies may confer protection against both depression and PTSD symptoms specifically in TBI exposed populations. Unlike the short-term effects of ketamine, however, natural anti-NMDAR1 autoantibody levels can persist for at least a year (**Supplemental Figure 1**) or longer and thereby provide long-term neuroprotection.

High titers of IgG isotype anti-NMDAR1 autoantibodies in brain contribute to the pathogenesis of anti-NMDAR1 encephalitis that exhibits a range of psychiatric symptoms^35^. We previously found that blood circulating IgG anti-NMDAR1 autoantibodies are sufficient to reduce spatial working memory in rodents with intact blood-brain barriers (BBB)^36^. Thus, the BBB may not be sufficient to prevent the entry of circulating anti-NMDAR1 autoantibodies into the brain, which is consistent with previous studies showing that ∼0.1% of circulating antibodies nonspecifically cross the BBB in healthy rodents and humans, regardless of antibody specificity or isotype^37–39^. In contrast to effects of IgG anti-NMDAR1 autoantibodies, our studies indicate that blood natural anti-NMDAR1 autoantibodies may confer protection against TBI-associated neuropsychiatric symptoms. How can these seemingly contradictory observations be reconciled? Natural autoantibodies in blood are predominantly the IgM isotype, with minor contributions from IgA and IgG3 ^40^. IgM anti-NMDAR1 autoantibodies may specifically inhibit extrasynaptic NMDARs and suppress glutamate excitotoxicity, but spare synaptic NMDARs, because IgM antibodies are physically too large to enter the synaptic cleft ^41^. Conceivably, suppression of glutamate excitotoxicity by natural anti-NMDAR1 autoantibodies, mostly IgM isotype, contributes to their neuroprotective effects against TBI-associated psychiatric symptoms. Unlike IgM, IgG anti-NMDAR1 autoantibodies are small enough to enter the synaptic cleft and inhibit synaptic NMDA neurotransmission, contributing to the pathological effects observed in anti-NMDAR1 encephalitis ^35^ and in immunized mice^36^.

We quantified total natural anti-NMDAR1 autoantibodies in blood, which are presumed to be predominantly of the IgM isotype^40^. Given potential presence of IgG or IgA anti-NMDAR1 autoantibodies in the quantification of total anti-NMDAR1 autoantibodies, neuroprotective effects of IgM anti-NMDAR1 autoantibodies may have been underestimated in our studies. Future studies should separately quantify IgM, IgG, and IgA isotypes to clarify their potentially divergent effects on psychiatric symptoms and cognitive function. A larger, independent cohort will be necessary to replicate neuroprotective effects of natural anti-NMDAR1 autoantibodies and IgM isotype, especially regarding nuances related to TBI severity, timing and type, for which our current sample size is limited. Many brain diseases and disorders involve some degree of glutamate excitotoxity. The neuroprotective potential of natural anti-NMDAR1 autoantibodies warrants investigation across a broader range of conditions beyond these current studies in TBI-associated depression and PTSD, Alzheimer’s disease^13^, and schizophrenia ^14,15^. Finally, IgM anti-NMDAR1 autoantibodies could be generated in *Aicda* mutant mice ^42^ via active immunization ^36^. Their neuroprotective effects could be investigated in mouse models. Success of these studies may open a new avenue for the development of the IgM antibody therapeutics for psychiatric disorders and many other brain diseases.

## Data Availability

All aggregate scientific data generated from this project will be made available as soon as possible under VHA Handbook 1200 guidelines, and no later than the time of publication or the end of the funding period, whichever comes first.

## Supplemental Figure Legend

**Supplemental Figure 1.**
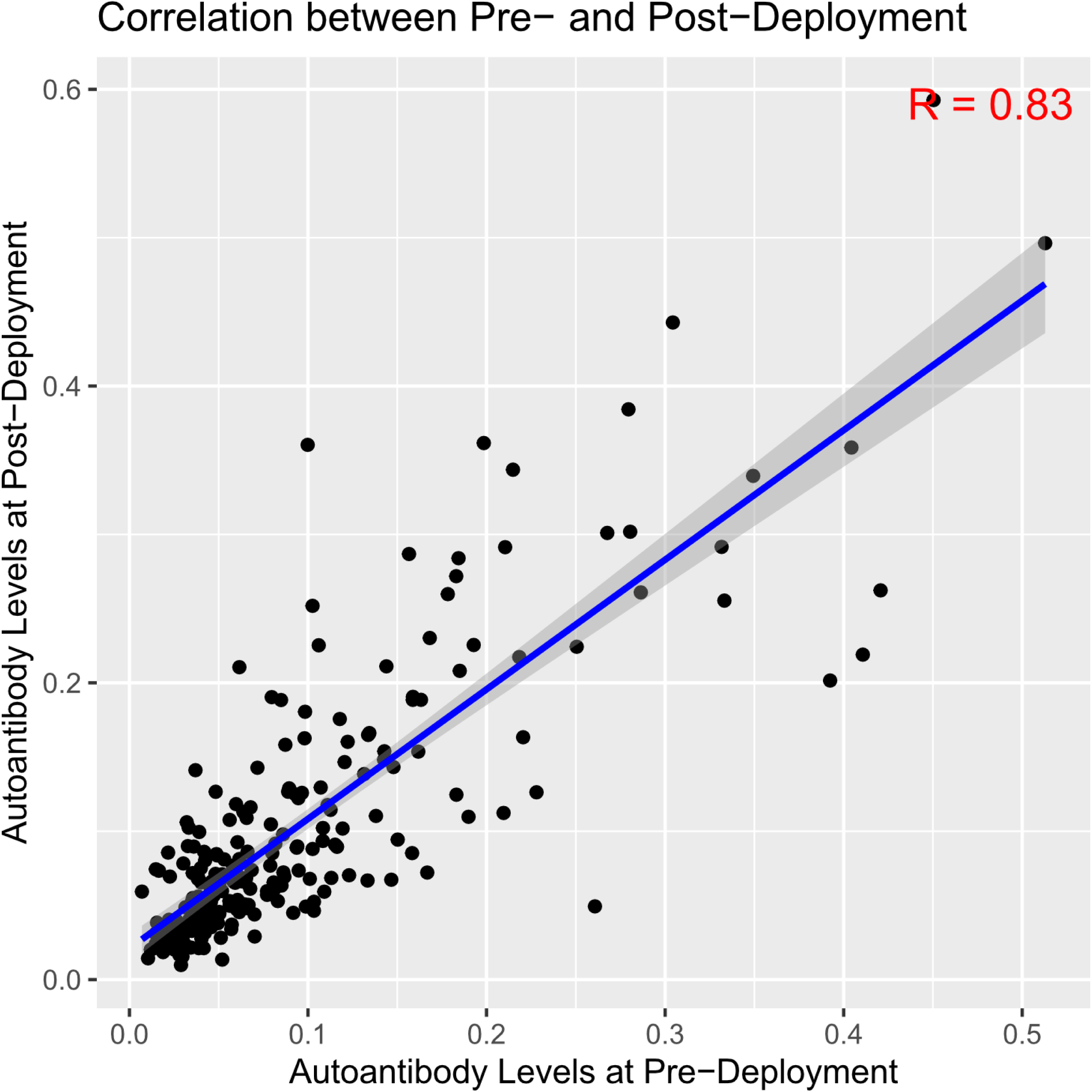
Correlation between pre-deployment and post-deployment levels of natural anti-NMDAR1 autoantibodies. A significant correlation was found with Pearson’s correlation analysis (R = 0.83, p < 0.001). Shaded regions represent the 95% confidence interval.

**Supplemental Figure 2.**
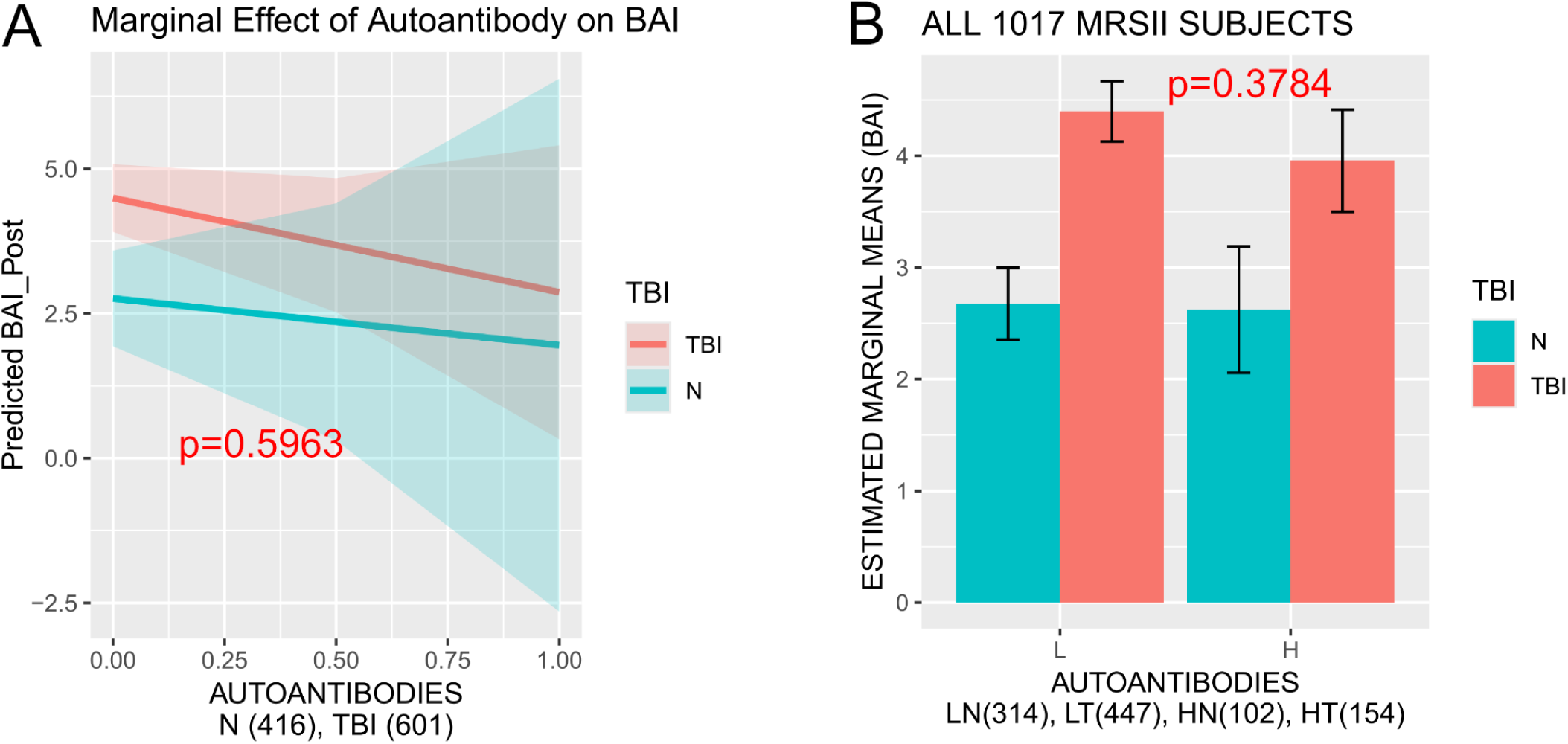
ANCOVA analysis of interaction between autoantibodies and TBI exposure on anxity. Legend: N = No TBI Exposure, TBI= TBI Exposure. Sample sizes (N) are indicated on the graphs. All models are visualized using non-transformed data, while statistical values are derived from linear models. P values < 0.10 are reported and color matched with TBI groups on the graphs. (**A)** Effect of continuous autoantibody levels on adjusted BAI anxiety scores. Shaded regions represent the 95% confidence interval. The regression model revealed a non-significant interaction between autoantibody levels and TBI (B= -0.265, p=0.6402). (**B**) Interaction between TBI and autoantibody levels as a categorical factor on adjusted BAI anxiety scores. A non-significant interaction was confirmed (F(1,1006) = 0.7764, p = 0.3784; FDR-adjusted p = 0.3784). Error bars = +\- SE.

**Supplemental Figure 3.**
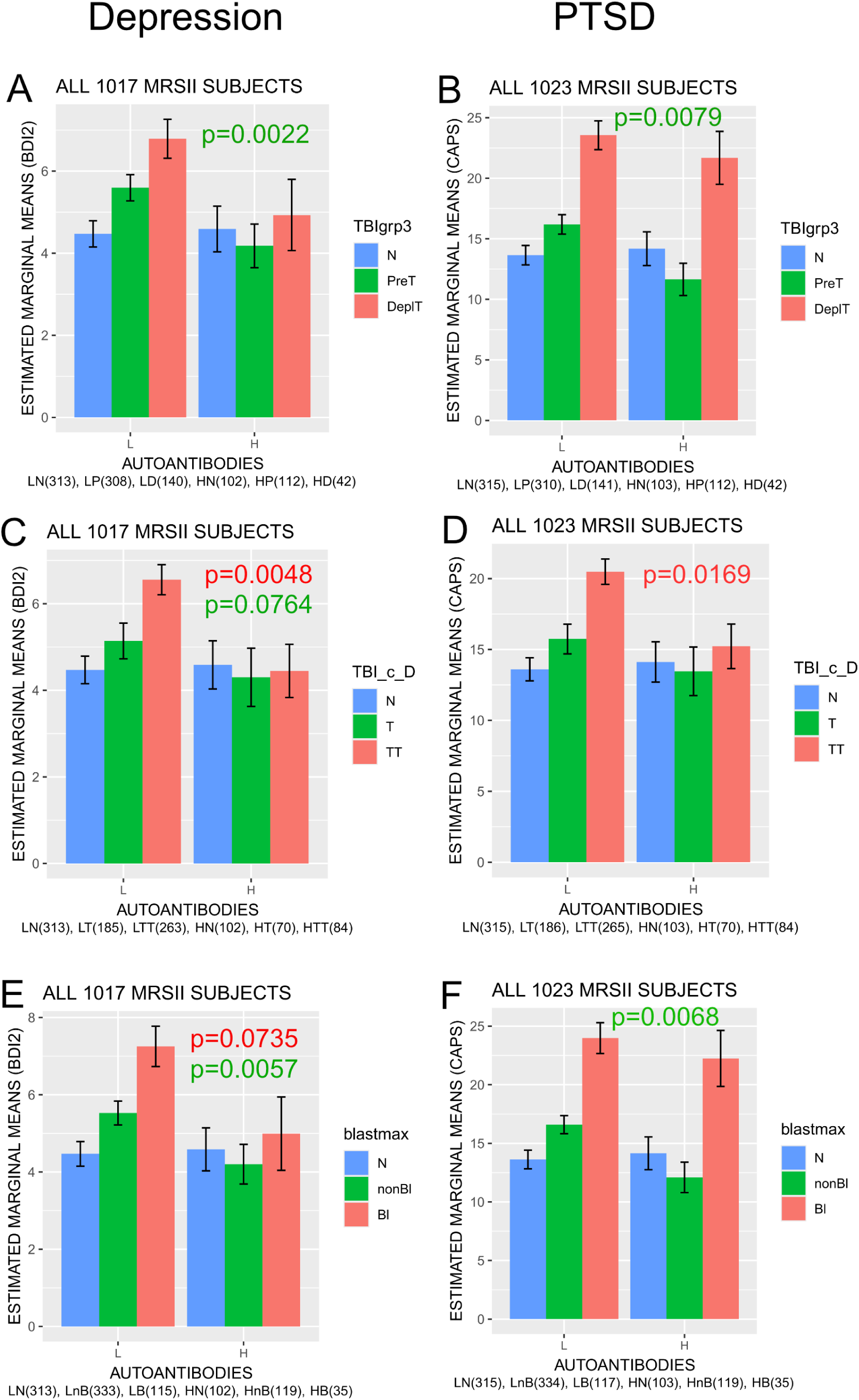
Secondary analyses evaluated the association between high anti-NMDAR1 autoantibody levels, treated as a categorical factor, and post-deployment depression and PTSD across TBI timing (A–B), number of injuries (C–D), and injury type (blast vs. non-blast; E–F). All models are visualized using non- transformed data, while statistical values are derived from linear models. P values < 0.10 are reported and color matched with TBI groups on the graphs. **Panel A-B** TBI timing: TBIgrp3 (TBI groups): N= No TBI Exposure, PreT= Pre-deployment TBI(s) only, DeplT= Deployment TBI. **(A)** Interaction between TBIgrp3 and autoantibody levels as a categorical factor on adjusted BDI2 depression scores. Significant interaction was confirmed (F(2,1008) = 4.3033, p = 0.0138; FDR-adjusted p = 0.0276). Pre-specified comparison between low and high autoantibody groups (p = 0.0022; Bonferroni corrected p = 0.0044), highlighting autoantibody protective effects against pre-deployment TBI. Error bars = +\- SE. **(B)** Interaction between TBIgrp3 and autoantibody levels as a categorical factor on adjusted CAPS PTSD scores. Significant interaction was confirmed (F(2,1015) = 3.0121, p = 0.0496; FDR-adjusted p = 0.0496). Pre-specified comparison between low and high autoantibody groups (p = 0.0079; Bonferroni corrected p = 0.0158), highlighting autoantibody protective effects against pre-deployment TBI. Error bars = +\-SE. **Panel C-D** Number of TBI injuries: TBI_c_D (TBI cumulative Deployment): N= No TBI Exposure, T= Single TBI, TT= Multiple TBIs. **(C)** Interaction between TBI_c_D and autoantibody levels as a categorical factor on adjusted BDI2 depression scores. Significant interaction was confirmed (F(2,1008) = 4.3191, p = 0.0136; FDR-adjusted p = 0.0272). Pre-specified comparison between low and high autoantibody groups (p = 0.0048; Bonferroni corrected p = 0.0096), highlighting autoantibody protective effects against multiple TBI. A trend toward significance was observed in single TBI (p = 0.0764; Bonferroni corrected p = 0.1528). Error bars = +\- SE. **(D)** Interaction between TBI_c_D and autoantibody levels as a categorical factor on adjusted CAPS PTSD scores. A trend toward significant interaction was confirmed (F(2,1015) = 2.8650, p = 0.0574; FDR- adjusted p = 0.0574). Pre-specified comparison between low and high autoantibody groups (p = 0.0169; Bonferroni corrected p = 0.0338), highlighting autoantibody protective effects against multiple TBI. Error bars = +\- SE. **Panel E-F** TBI Type: blastmax: N= No TBI Exposure, nonBl= Non-blast TBI only, Bl= Blast TBI. **(E)** Interaction between blastmax and autoantibody levels as a categorical factor on adjusted BDI2 depression scores. Significant interaction was confirmed (F(2,1008) = 4.1922, p = 0.0154; FDR-adjusted p = 0.0308). Pre-specified comparison between low and high autoantibody groups (p = 0.0057; Bonferroni corrected p = 0.0114), highlighting autoantibody protective effects against non-blast TBI. A trend toward significance was observed in blast TBI (p = 0.0735; Bonferroni corrected p = 0.1470). Error bars = +\- SE. **(F)** Interaction between blastmax and autoantibody levels as a categorical factor on adjusted CAPS PTSD scores. Significant interaction was confirmed (F(2,1015) = 3.0677, p = 0.0469; FDR-adjusted p = 0.0469). Pre-specified comparison between low and high autoantibody groups (p = 0.0068; Bonferroni corrected p = 0.0136), highlighting autoantibody protective effects against non-blast TBI. Error bars = +\- SE.

**Supplemental Figure 4.**
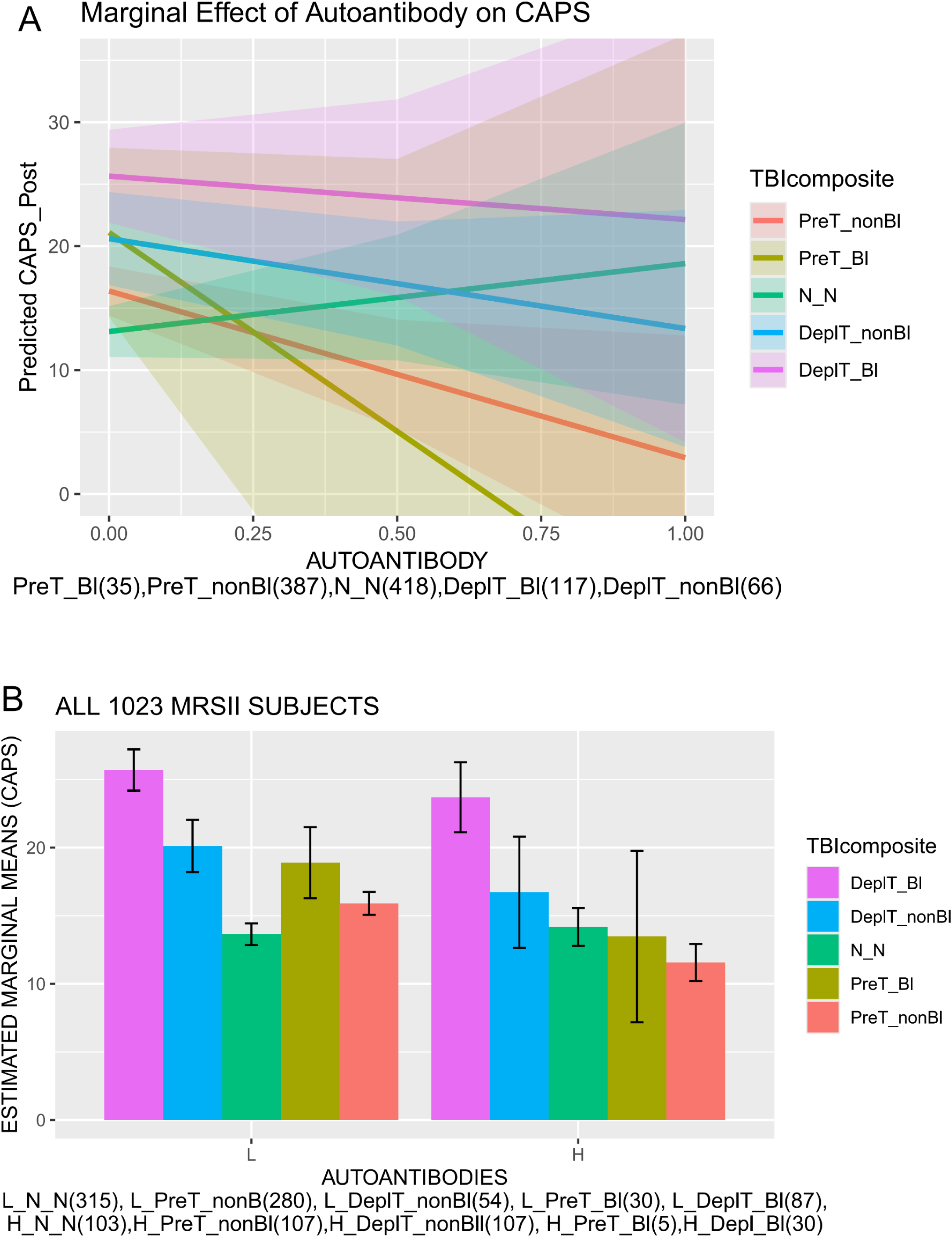
Association of high levels of anti-NMDAR1 autoantibody levels with reduced scores of psychiatric symptoms in subjects with blast TBI occurred during pre-deployment. TBIcomposite: N = No TBI Exposure, PreT_nonBl=Pre-deployment TBI only with non-blast TBI type. PreT_Bl=Pre-deployment only with blast TBI, DeplT_nonBl= Deployment TBI with non-blast TBI, DeplT_Bl=Deployment TBI with blast TBI. Sample sizes (N) are indicated on the graphs. All models are visualized using non-transformed data, while statistical values are derived from linear models. P values < 0.10 are reported and color matched with TBI groups on the graphs. (**A)** Effect of continuous autoantibody levels on adjusted CAPS-IV scores. Shaded regions represent the 95% confidence interval. The regression model revealed a non-significant interaction between autoantibody levels and PreT_Bl (B= -2.16, p = 0.4151). (**B**) A significant TBI effect was observed (F(4, 1011)=11.9998, p=1.569e-09). Interaction between TBIcomposite and autoantibody levels as a categorical factor on adjusted CAPS PTSD scores. Direct comparison between low and high autoantibody groups with pre-deployment blast TBI (PreT_Bl) revealed a non-significant effect (p = 0.6851), likely due to small sample sizes. Error bars = +\- SE.

**Supplemental Table 1.** Demographics by Autoantibody as a Categorical Factor.

|  | Total Cohort<br>(n=1025) | Low<br>(n=768) | High<br>(n=257) | Test Statistic | P Value |
| --- | --- | --- | --- | --- | --- |
| **Age (Years)** |  |  |  |  |  |
| Mean(SD) | 21.9 (2.7) | 21.9 (2.7) | 21.7 (2.7) |  |  |
| Range | 18-42 | 18-37 | 18-42 | Z=-1.225 | p=0.221 |
| **Education (Years)** |  |  |  |  |  |
| Mean (SD) | 12.3 (0.87) | 12.3 (0.81) | 12.3 (0.96) | Z=-.092 | p=0.927 |
| Range | 10--20 | 10--18 | 11--20 |  |  |
| **Genetic Ancestry, N(%)** | | | | $\chi^2=5.572(df=3)$ | p=0.134 |
| African | 43 (4.2%) | 31(4.0%) | 12 (4.7%) |  |  |
| European | 553 (54%) | 437 (56.9%) | 116 (45.1%) |  |  |
| Hispanic/Native American | 169 (16.5%) | 123 (16.0%) | 46(17.9%) |  |  |
| Asian/Other | 99 (9.7%) | 70 (9.1%) | 29 (11.3%) |  |  |
| Missing | 161 (15.7%) | 107 (13.9%) | 54 (21%) |  |  |
| **Deployment Status** | | | | $\chi^2=4.537(df=1)$ | **p=0.033\*** |
| Yes | 850 (82.9%) | 648 (84.4%) | 202 (78.6%) |  |  |
| No | 175 (17.1%) | 120 (15.6%) | 55 (21.4%) |  |  |
| **Prior Deployment** | | | | $\chi^2=4.909(df=1)$ | **p=0.027\*** |
| Yes | 508 (49.6%) | 396 (51.6%) | 112 (43.6%) |  |  |
| No | 517 (50.4%) | 372 (48.4%) | 145 (56.4%) |  |  |
| **TBI Status** | | | | $\chi^2=.937(df=2)$ | p=.626 |
| No TBI | 419 (40.9%) | 316 (41.1%) | 103 (40.1%) |  |  |
| Predeployment TBI only | 423 (41.3%) | 311 (40.5%) | 112 (43.6%) |  |  |
| Deployment TBI | 183 (17.9%) | 141 (18.4%) | 42 (16.3%) |  |  |
| **Cumulative # TBIs** | | | | $\chi^2=.971(df=2)$ | p=0.615 |
| 0 | 419 (40.9%) | 316 (41.2%) | 103 (40.1%) |  |  |
| 1 | 256 (25%) | 186 (24.2%) | 70 (27.2%) |  |  |
| 2+ | 350 (34.1%) | 266 (34.6%) | 84 (32.7%) |  |  |
| **TBI Type** | | | | $\chi^2=.706(df=2)$ | p=.703 |
| Nonblast | 454 (44.3%) | 335 (43.6%) | 119 (46.3%) |  |  |
| Blast | 152 (14.8%) | 117 (15.2%) | 35(13.6%) |  |  |
| **DRRI Composite** |  |  |  | T=.682<br>(df=846) | p=.496 |
| Mean (SD) | .0148 (1.77) | .0379 (1.8) | -.0596(1.68) |  |  |
| Range | -3.11-7.23 | -3.11-7.23 | -3.11-6.54 |  |  |
| 2+ | 350 (34.1%) | 266 (34.6%) | 84 (32.7%) |  |  |
| **Psychotropic Medication @ Post-deployment** | | | | $\chi^2=7.594(df=1)$ | **p=0.006** |
| Yes | 36 (3.5%) | 34 (4.4%) | 2 (0.8%) |  |  |
| No | 987 (96.3%) | 732 (95.3%) | 255 (99.2%) |  |  |
| Missing | 2 (0.2%) | 2 (0.2%) |  |  |  |
| **Psychotropic Medication @ Pre-deployment** | | | | $\chi^2=0.54(df=1)$ | p=0.462 |
| Yes | 8 (0.8%) | 7 (0.9%) | 1 (0.4%) |  |  |
| No | 853 (83.2%) | 652 (84.9%) | 201 (78.2%) |  |  |
| missing | 164 (16%) | 109 (14.2%) | 55 (21.4%) |  |  |

**Supplemental Table 2:** Simple Slopes of continuous multiple linear regression. Trending or significant p values are bolded (p<0.10). SE=standard error. CI = 95% confidence interval.

| Dependent Variable | Moderator | Level | Effect | SE | t -value | p-value | CI (95%) |
| --- | --- | --- | --- | --- | --- | --- | --- |
| **BDI-II**<br>**(Depression)** | Lifetime TBI Exposure | No TBI | 0.3421 | 0.4155 | 0.8233 | 0.4105 | [-.4733, 1.1574] |
|  |  | TBI Exposure | -1.1967 | 0.3037 | -3.9408 | **0.0001** | [-1.7926, -.6008] |
|  | Deployment TBI History | No TBI | .3382 | .4144 | .8161 | .4146 | [-.4750, 1.1514] |
|  |  | Pre-deployment TBI | -1.2734 | .3759 | -3.3880 | **.0007** | [-2.0110, -.5359] |
|  |  | Deployment TBI | -1.2043 | .5147 | -2.3397 | **.0195** | [-2.2144, -.1943] |
|  | Cumulative # TBIs | No TBI | 0.3398 | 0.4145 | 0.8199 | 0.4124 | [-.4735, 1.1532] |
|  |  | Single TBI | -1.1022 | 0.4274 | -2.5792 | **0.0100** | [-1.9409, -.2636] |
|  |  | Multiple TBI | -1.2457 | 0.4305 | -2.8935 | **0.0039** | [-2.0904, -.4009] |
|  | TBI Type | No TBI | 0.3389 | 0.4134 | 0.8198 | 0.4125 | [-.4723, 1.1501] |
|  |  | Non-Blast TBI | -1.28 | 0.3415 | -3.7484 | **0.0002** | [-1.9502, -.6099] |
|  |  | Blast TBI | -0.9197 | 0.652 | -1.4106 | 0.1587 | [-2.1991, .3597] |
| **CAPS**<br>**(PTSD)** | Lifetime TBI Exposure | No TBI | 0.9626 | 0.6631 | 1.4516 | 0.1469 | [-.3386, 2.2638] |
|  |  | TBI | -0.9521 | 0.4877 | -1.9522 | **0.0512** | [-1.9091, .0049] |
|  | Deployment TBI History | No TBI | 0.9501 | 0.6512 | 1.4591 | 0.1449 | [-.3277, 2.2279] |
|  |  | Pre-deployment TBI | -1.2208 | 0.5942 | -2.0544 | **0.0402** | [-2.3869, -.0547] |
|  |  | Deployment TBI | -1.005 | 0.8146 | -1.2337 | 0.2176 | [-2.6034, .5935] |
|  | Cumulative # TBIs | No TBI | 0.9556 | 0.6611 | 1.4456 | 0.1486 | [-.3416, 2.2529] |
|  |  | Single TBI | -0.8443 | 0.6862 | -1.2304 | 0.2189 | [-2.1909, .5023] |
|  |  | Multiple TBI | -0.9779 | 0.6904 | -1.4165 | 0.1569 | [-2.3327, .3768] |
|  | TBI Type | No TBI | 0.9545 | 0.6534 | 1.4607 | 0.1444 | [-.3278, 2.2367] |
|  |  | Non-Blast TBI | -1.1258 | 0.5435 | -2.0714 | **0.0386** | [-2.1922, -.0593] |
|  |  | Blast TBI | -0.3665 | 1.0371 | -0.3534 | 0.7329 | [-2.4017, 1.6686] |

**Supplemental Table 3.** Effect Sizes.

| **TBI Category** | **Dependent Variables** | **Group Size L (n1)** | **Group Size H (n2)** | **Effect Size (Cohen's d)** |
| --- | --- | --- | --- | --- |
| **Lifetime TBI (TBI exposure)** | CAPS | 451 | 154 | 0.25 |
|  | BDI2 | 448 | 154 | 0.32 |
| **TBIgrp3 (PreDeployment TBI)** | CAPS | 310 | 112 | 0.29 |
|  | BDI2 | 308 | 112 | 0.34 |
| **TBI_c_D (Multiple TBI)** | CAPS | 265 | 84 | 0.3 |
|  | BDI2 | 263 | 84 | 0.36 |
| **TBI Type (Non-Blast TBI)** | CAPS | 334 | 119 | 0.29 |
|  | BDI2 | 333 | 119 | 0.3 |

